# Implications of the reverse associations between obesity prevalence and coronavirus disease (COVID-19) cases and related deaths in the United States

**DOI:** 10.1101/2020.06.09.20127035

**Authors:** Ning Zhang, Lorraine S. Cordeiro, Zhenhua Liu

## Abstract

The Coronavirus disease 2019 (COVID-19), caused by severe acute respiratory syndrome coronavirus 2 (SARS-CoV-2) that was most recently discovered, quickly evolved into a global pandemic. Studies suggested that obesity was a major risk factor for its hospitalization and severity of symptoms. This study investigated the associations between obesity prevalence with overall COVID-19 cases and related deaths across states in the United States. General regression and Chi-square tests were used to examine those associations. The analyses indicated that obesity prevalence (%) across states were negatively associated with COVID-19 cases (*p* = 0.0448) and related deaths (*p* = 0.0181), with a decrease of 158 cases/100K population and 13 deaths/100K for every 5% increase of the obesity prevalence. When the states were divided based on the median of obesity prevalence (30.9%) into a group of states with low obesity prevalence and a group with high obesity prevalence, both the cases (671 vs 416 cases/100k population) and deaths (39 vs 21 deaths/100k population) were significantly different (*p* < 0.001) across groups. These findings provided important information for the relationship between the dual pandemic threats of obesity and COVID-19. These results should not currently be considered as an indication that obesity is a protective factor for COVID-19, and would rather be used as a warning of the public advice that obese people is more vulnerable to COVID-19 infection, which may lead to a false safety message probably given to people with normal body weight.

## Introduction

The Coronavirus disease 2019 (COVID-19) is a new infectious disease most recently discovered with the first case reported in December, 2019 ^1^. It was resulted from the infection of a novel coronavirus called severe acute respiratory syndrome coronavirus 2 (SARS-CoV-2) and was named as COVID-19 by the World Health Organization (WHO) on March 11, 2020. It is now classified as a global pandemic ^2^. As of May 31, 2020, there were ∼6 million cases and ∼371,000 related deaths worldwide, with ∼1,788,000 cases and over 100,000 deaths in the United States alone ^3^. COVID-19 represents the third epidemic of coronavirus infections in the 21^st^ century following severe acute respiratory syndrome (SARS)-CoV (2002) ^4^ and the Middle East respiratory syndrome (MERS)-CoV (2012) ^5^. The most common symptom of COVID-19 is fever, often accompanied by other symptoms including but not limited to cough, loss of appetite, fatigue, shortness of breath, and sputum production ^6–8^.

Obesity, defined as BMI of 30 kg/m^2^ or greater, has experienced an unprecedented rise in recent decades in the United States ^9, 10^ and also in many other industrialized and urban areas in developing countries throughout the world ^11, 12^. It has reached an epidemic level in US and a further increase, from the current 1/3 of the population to ∼50% by 2030, is projected ^13, 14^. Obesity is associated with increased risks of developing various health problems, including type 2 diabetes mellitus (T2M), cardiovascular diseases and stroke, hypertension and various cancers ^15, 16^. Most people with obesity show signs of a low-grade, chronic inflammation ^17^, which plays as a critical underlying mechanism responsible for those obesity-associated medical complications ^18^.

Studies have shown that COVID-19 poses a particular risk to people living with preexisting complications. Data from a study in China indicated that the fatality of patients with a lab-confirmed COVID-19 is ∼2.3%, but the number increased to 10.5% for patients with cardiovascular disease, 7.3%t for those with diabetes, 6.3% for those with chronic respiratory disease, 6.0% for those with hypertension, and 5.6% for those with cancer ^19^. As these health conditions are directly associated with obesity ^15, 16^, it is not surprising that many recent studies show that obesity is associated with more severe symptoms and increased intensive care admissions of COVID-19 patients ^20–24^.

Most recent studies showed that obesity, especially morbid obesity with BMI ≥ 40 kg/m^2^, is an independent risk factor of hospitalization, severity of symptoms and intensive care admissions of COVID-19 ^25^. However, this apparent evidence contradicts with the “obesity paradox” theory - obesity and morbid obesity were associated with a lower mortality rate in patients with acute respiratory distress syndrome ^26, 27^. The current study examined how obesity prevalence influence the overall infection of SARS-CoV-2 and COVID-19 related deaths at a population level. Our analysis demonstrated negative associations between obesity prevalence and coronavirus disease (COVID-19) cases and related deaths in the United States. These results provided interesting and important information for policy makers to combat the pandemic of COVID-19.

## Materials and methods

We examined the incidence and mortality data in the United States from three different resources: the COVIDTracking Project (COVIDTracking, https://covidtracking.com), the US Centers for Disease Control and Prevention (CDC, https://www.cdc.gov/coronavirus/2019-nCoV), and The Coronavirus Resource Center at John Hopkins University (JNU, https://coronavirus.jhu.edu). We used the data retrieved from these resources on May 31, 2020, which covers the cases and deaths over the course of more than 4 months from January 22, 2020 when the first case was identified according to the CDC. The population data for each state was the estimated population by July 1, 2019 and was released by the U.S. Census Bureau, Population Division (https://www.census.gov).

We focused on data from 50 states plus the District of Columbia without including other US territories. Statistical analyses were carried out using general linear regression and Chi-square tests. Four different regression models were performed to examine the associations between obesity prevalence and COVID-19 cases and related deaths. Our first model examined the crude associations between obesity prevalence and COVID-19 cases and deaths without adjusting for any confounding factors. Since our most recent study ^28^, as well as other studies ^29, 30^, indicated that sunlight and low vitamin D levels are possible risk factors for COVID-19, our second model adjusted for a geographic factor, using latitude as an index. With consideration of the significant variations of COVID-19 testing rates across the states, our third model also included the test rate as confounding factor. With further consideration of population density, especially New York City as the epicenter of COVID-19 in US, we further adjusted for population in our fourth model. We adjusted for population rather the population density as we realize that even though New York City has a high population density, but the population density for the whole state is not high. Instead, New York is among the states with a high population, therefore using the population as confounding factor is appropriate. All data are presented in the Supplementary File (Table S1). Statistical analyses were performed using SAS program (Version 9.4, SAS Institute, Cary, NC, USA).

## Results

### Characteristics of the population

As of May 31, 2020, there was a total of 1,787,680 COVID-19 cases after the first case was reported in the US on Jan 22, 2020 (CDC, Fig. 1A). Obesity prevalence ranged from 23.0% to 39.5% with a mean of 31.3 ± 0.5% across the states (Fig. 1B). The overall rates of COVID-19 cases and related deaths are 542 cases/100K population and 30 deaths/100K population, respectively (Fig. 1C and 1D).

**Figure 1.**
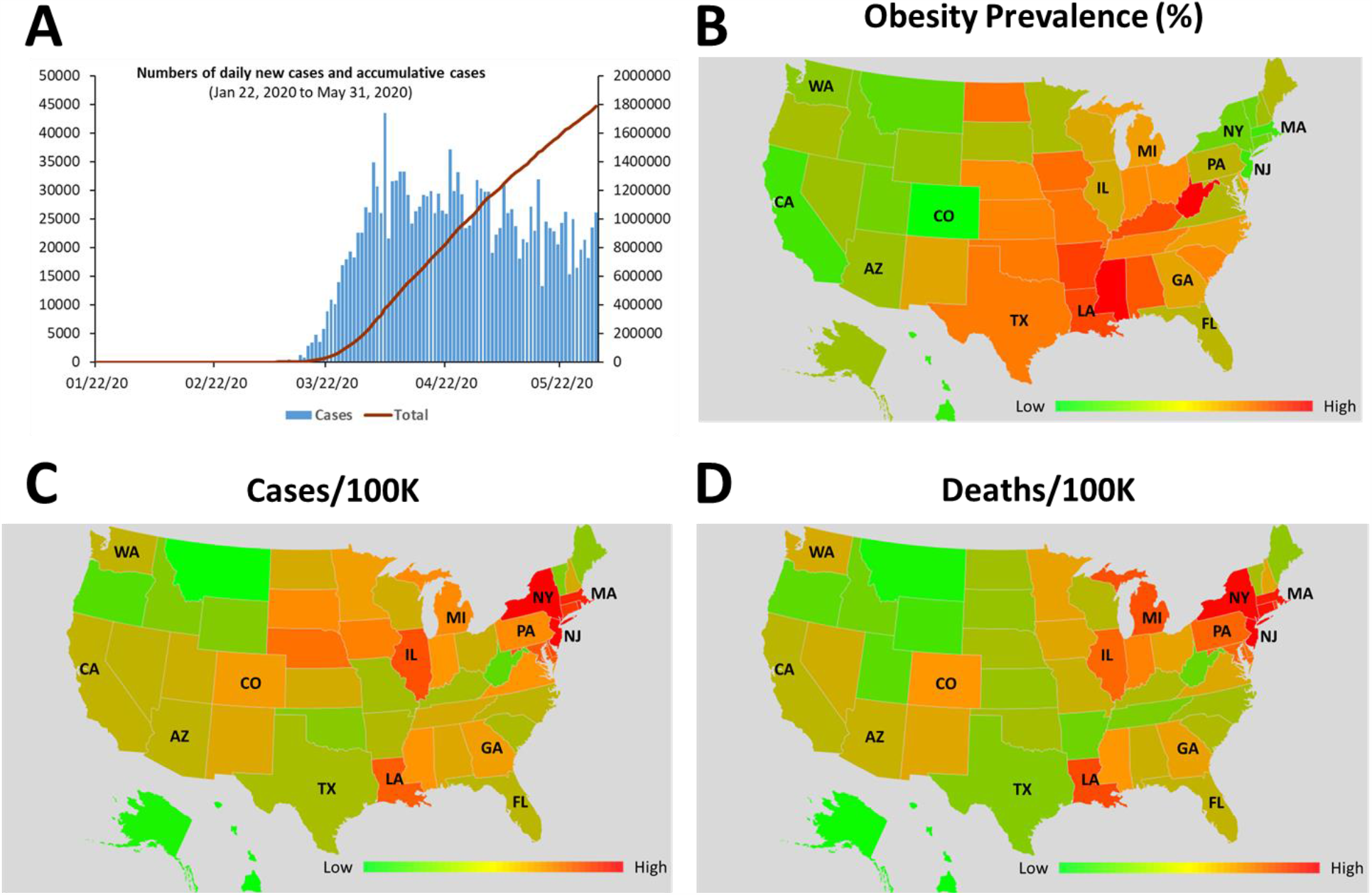
Characteristics of the population. A) By May 31, 2020, there are a total of 1,787,680 cases after the first case reported on Jan 22, 2020 (data from CDC); B) Obesity prevalence (%) across states; C) COVID-19 cases (cases/100K population) across states; D) COVID-19-related deaths (deaths/100K population) across states.

### Correlations between obesity prevalence and COVID-19 cases and related deaths

The results from all four different regression models revealed that obesity prevalence across states was negatively associated with both COVID-19 cases (Table 1) and related deaths (Table 2) regardless of adjustments made for the three confounding factors. Significant reverse associations were observed for COVID-19 related deaths in all 4 models (*p* < 0.05), and significant associations were also observed for COVID-19 cases in the crude model, Model I and II (*p* < 0.05) with a marginally significant association observed in Model III (*p* = 0.0645). According to the crude model, every 5% increase of obesity prevalence was associated with a decrease of 158 cases/100K population (Fig. 2A) and 13 deaths/100K population (Fig.2B) (*p* < 0.05).

**Table 1.** The regression between COVID-19 cases per 100K population (by May 31, 2020) and obesity prevalence (%) across states in the United States.

| **Models** | **β-Coefficient<sup>5</sup>**<br>for obesity prevalence (%) | **Cases of Changes<sup>6</sup>**<br>for every 5% of<br>obesity prevalence | ***p*-value** |
| --- | --- | --- | --- |
| **Crude Model<sup>1</sup>** | -31.69 ± 15.39 | -158.45 | 0.0448 |
| **Model I<sup>2</sup>** | -32.11 ± 15.95 | -160.55 | 0.0497 |
| **Model II<sup>3</sup>** | -24.69 ± 11.93 | -123.45 | 0.0441 |
| **Model III<sup>4</sup>** | -22.89 ± 12.09 | -114.45 | 0.0645 |

<sup>1</sup> Crude model: Without adjustment.
<sup>2</sup> Model I: Adjusted for latitude.
<sup>3</sup> Model II: Adjusted for both latitude and test rates in each state.
<sup>4</sup> Model III: Adjusted for latitude, test rates and population in each state.
<sup>5</sup> Values are regression $\beta$ -coefficients ± SEs, based on the COVID-19 cases/100K population in each states.
<sup>6</sup> Values are the number of case changes for each 5% increase of the obesity prevalence.

**Table 2.** The regression between COVID-19 related deaths per 100K population (by May 31, 2020) and obesity prevalence (%) across states in the United States.

| **Models** | **β-Coefficient<sup>5</sup>**<br>for obesity prevalence (%) | **Cases of Changes<sup>6</sup>**<br>for every 5% of<br>obesity prevalence | ***p*-value** |
| --- | --- | --- | --- |
| **Crude Model<sup>1</sup>** | -2.68 ± 1.10 | -13.40 | 0.0181 |
| **Model I<sup>2</sup>** | -2.71 ± 1.14 | -13.55 | 0.0209 |
| **Model II<sup>3</sup>** | -2.25 ± 0.93 | -11.25 | 0.0192 |
| **Model III<sup>4</sup>** | -2.07 ± 0.93 | -10.35 | 0.0310 |

<sup>1</sup> Crude model: Without adjustment.
<sup>2</sup> Model I: Adjusted for latitude.
<sup>3</sup> Model II: Adjusted for both latitude and test rates in each state.
<sup>4</sup> Model III: Adjusted for latitude, test rates and population in each state.
<sup>5</sup> Values are regression $\beta$ -coefficients ± SEs, based on the COVID-19 related deaths/100K population in each states.
<sup>6</sup> Values are the number of death changes for each 5% increase of the obesity prevalence.

**Figure 2.**
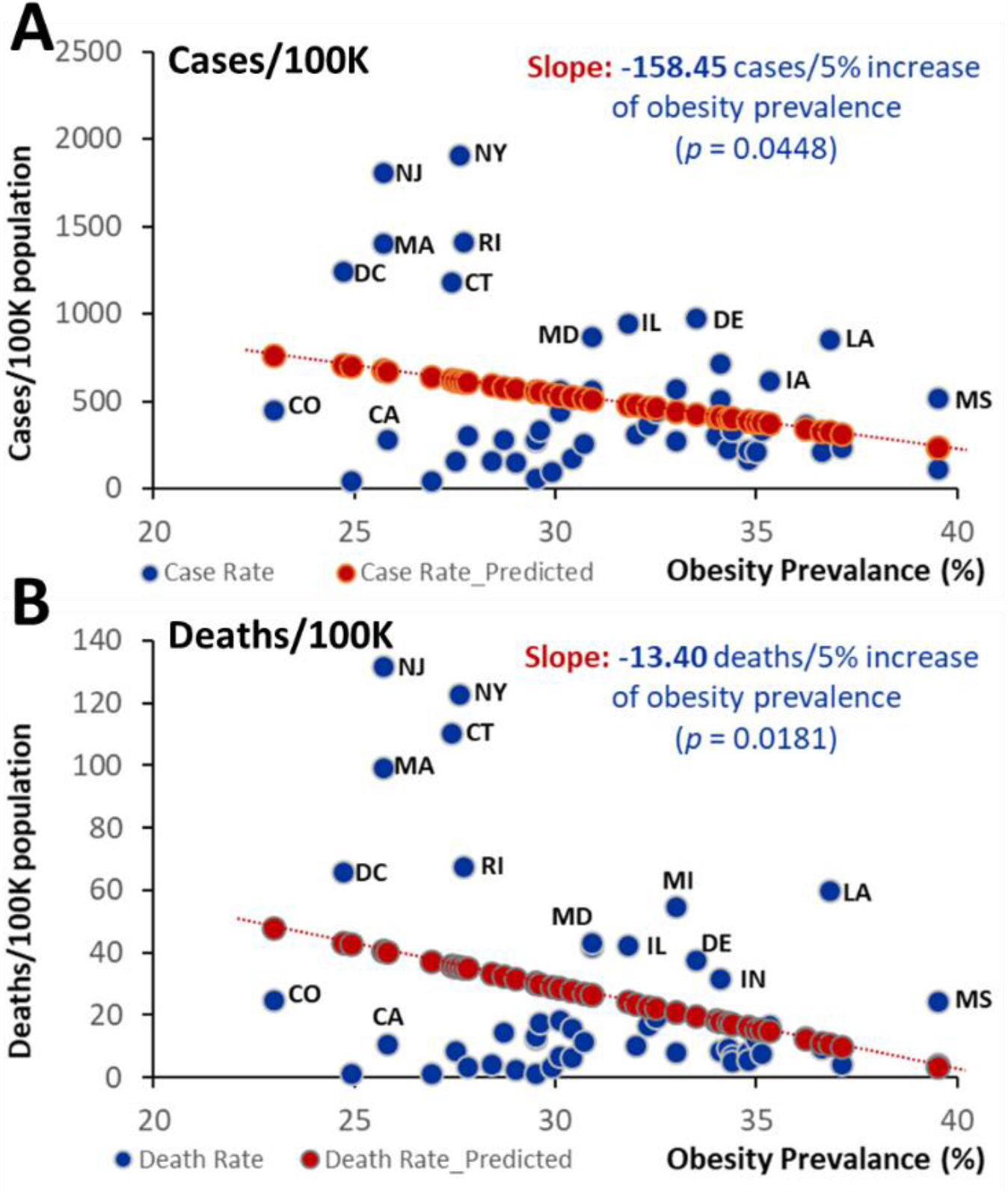
Correlations between obesity prevalence (%) and COVID-19 cases and related deaths. A) Cases. Every 5% increase of obesity prevalence represents a decrease of 158 cases/100K population of COVID-19 cases, B) Deaths. Every 5% increase of obesity prevalence represents a decrease of 13 deaths/100K population of COVID-19 related deaths in the United States.

Among the states with cases more than 1000 cases/100K population, five states (i.e. New York, New Jersey, Massachusetts, Connecticut, and Rhode Islands), as well as the District of Columbia, had an obesity prevalence below the median (30.9%). Among the states with mortality greater than 50 deaths/100K population, five states (New York, New Jersey, Massachusetts, Connecticut, and Rhode Islands) as well as D.C. has an obesity prevalence below the median (30.9%), and only two states (i.e. Michigan and Louisiana) had an obesity prevalence above the median (30.9%) (Fig. 2A and 2B).

### Comparisons of COVID-19 cases and related deaths between the states with obesity prevalence above and below the median (30.9%)

We categorized states into two groups for comparison: one group with obesity prevalence below the median (30.9%) (OP_Low) and the other group with obese prevalence above the median (30.9%) (OP_High). Chi-square tests were performed to compare COVID-19 cases and related deaths between the two groups. Our analyses suggested that states with obesity prevalence above the median (≥ 30.9%) had significantly lower COVID-19 cases (416 vs 671 cases/100k population) and related deaths (21 vs 39 deaths/100k population) than states with obesity prevalence below the median (30.9%) (*p* < 0.001) (Fig. 3A and 3B).

**Figure 3.**
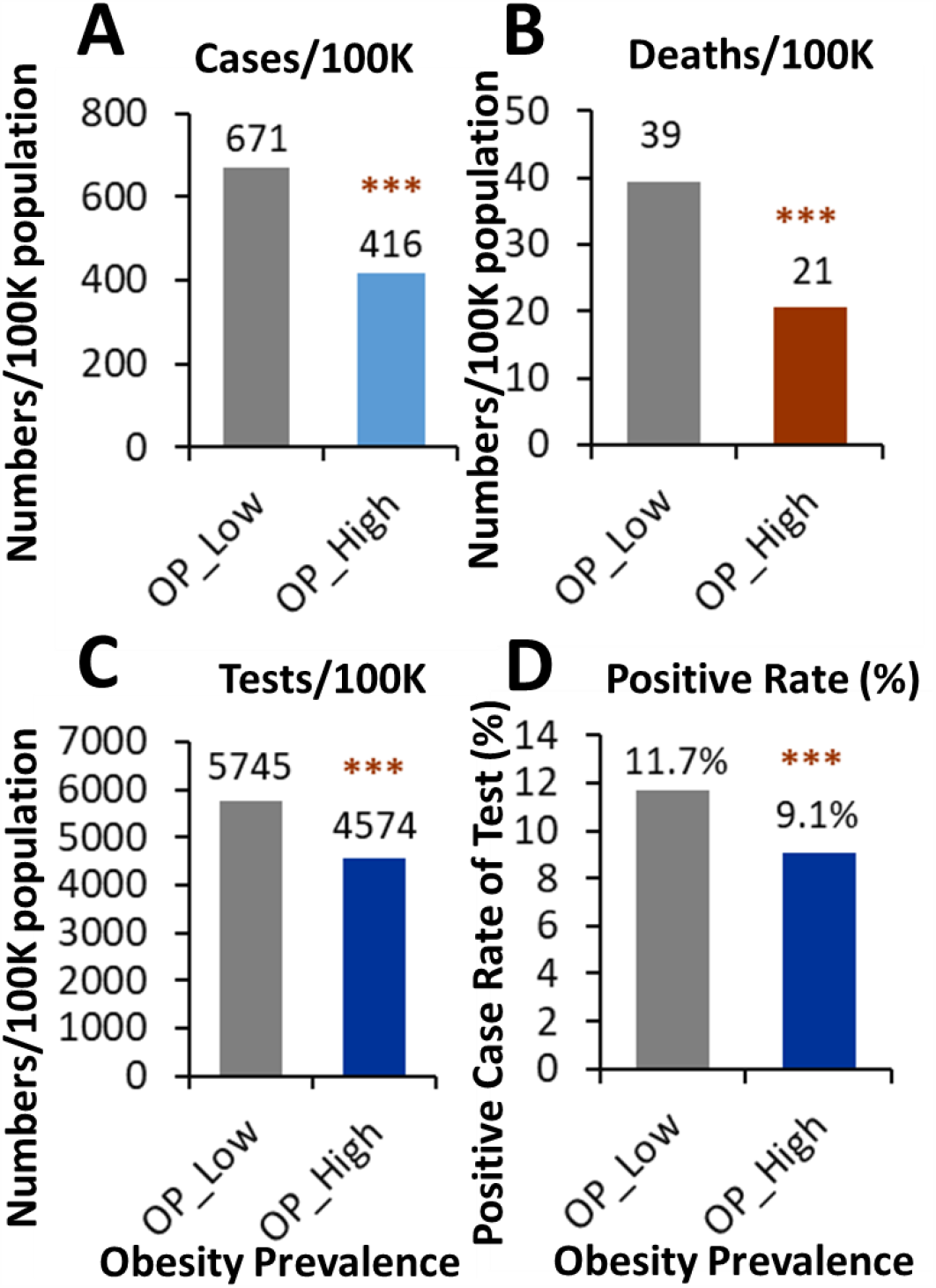
COVID-19 cases and related deaths separated by the median of obesity prevalence. A) COVID-19 cases (cases/100K population); B) COVID-19 mortalities (deaths/100K population); C) Test rate (numbers/100K population). How many individuals were tested for COVID-19 per 100K population; D) Positive case rate (%). The percentage of positive case in the subject tested for COVID-19. OP_Low and OP_High: the obesity prevalence low than the median or high than the median (30.9%).

It is noted that the states with high obesity prevalence had lower COVID-19 testing rates than states with low obesity prevalence (4574 vs 5745 individuals/100K population) (Fig.3C). However, the significantly higher levels of COVID-19 cases and related deaths per 100K population in the states with low obesity prevalence were not attributed to the high testing rate, as positive rate of testing in the states with low obesity prevalence were even higher (*p* < 0.01) than those states with high obesity prevalence (11.7% vs 9.1%) (Fig. 3D). The high positive rate over the total tested indicates COVID-19 was more prevalent in the states with low obesity prevalence than those states with high obesity prevalence (Fig. 3D).

## Discussion

Using the accumulated COVID-19 cases (over 1,788,000) and related deaths (over 104,000) data from the first case reported by CDC on Jan 22, up to May 31, 2020, we surprisingly found reversed associations between obesity prevalence and COVID-19 cases and associated mortalities in 50 states plus DC in the United States.

When COVID-19 cases and related deaths across different states were plotted against obesity prevalence in each state regardless of adjusting for the geographic factor (latitude), testing rates or population size, we found that obese prevalence was negatively associated with both COVID-19 cases and related deaths, with a decrease of 158 cases/100K population and 13 related deaths/100K population per 5% increase of the obesity prevalence (*p* < 0.05). When the states were divided into high and low obesity prevalence groups based on the median threshold of the obesity prevalence (30.9%), both the COIVD-19 cases and related deaths were significantly higher (*p* < 0.001) in the states with low obesity prevalence, when compared to the states with high obesity prevalence.

Most recent studies however reported that obesity is a risk factor for the hospitalization, severity of symptoms and intensive care admissions of COVID-19 ^20–22^, and it is particularly the case for young to middle age people under 60 years of age ^23, 24^. These studies have led to the public advices that obese people, especially those with BMI ≥ 40 kg/m^2^, are more vulnerable to COVID-19, comparing to people with normal BMI < 25 kg/m^2^ ^20, 31^. However, there are also publications reporting that there are lack of clarity for this guidance of obesity and COVID-19 ^26,32^. Previous studies showed that obesity was associated with a decrease of mortality in patients with acute respiratory distress syndrome and it was referred to as the “obesity paradox” ^27, 33^. The claim that obesity is an independent risk factor for COVID-19 is apparently disagreement with the “obesity paradox” theory ^26^. The pathophysiological mechanism underlying this theory is that obesity represents a preconditioned chronic pro-inflammatory status, which creates a protective environment that limited the detrimental effects from a more aggressive second hit ^26, 34^.

The present study reported a negative association between obesity prevalence and COVID-19 outbreak. Our study focused on how obesity prevalence influence the possibility of overall infection and lethality of COVID-19 at a population level. It is different from the aforementioned studies that mainly focused on the influence of obesity on the hospitalization, severity of symptoms and clinical outcomes. To our knowledge, our findings represent the first report showing a potential negative relationship between obesity prevalence and COVID-19 cases and related deaths. These findings, though counterintuitive, are mechanistically in agreement with the aforementioned “obesity paradox” theory. In addition, these results may also reflect the influence of the lifestyle associated with obesity people. One of the major contributions for obesity is sedentary lifestyle with limited out-door exercise ^35^, which is protective for the highly transmissible COVID-19.

We acknowledge that this study has limitations: a) There are large variations of the COVID-19 cases and related mortalities across the states, which reduced the accuracy of the estimate. b) The number of cases/state is heavily affected by the number of tests, which are unequally performed across the states. However, even with highly varied test rates across states, the higher test rate/state (5745 vs 4574/100K, Fig. 3C) together with a higher positive rate/tests (11.7 vs 9.1%, Fig. 3D) for the states with low obesity prevalence collectively suggested the higher prevalence of COVID-19 in those states, comparing to states with high obesity prevalence. Also, even after adjusting for the test rate, the results remains significant (Model II). c) COVID-19 is a highly transmissible disease, and population density in large cities are a critical risk factor. Even though in our model III we adjusted for the population, we recognized the influence from the New York City as the epicenter. Nevertheless, the present study discovered interesting negative associations between obesity prevalence and COVID-19 cases and related deaths. These findings may possibly be explained by the “obesity paradox” theory and the sedentary lifestyle with limited out-door activity that is potentially protective to COVID-19 infection in obese people.

In conclusion, the present study observed interesting negative associations between obesity prevalence with COVID-19 cases and related mortalities. These associations requires further mechanistic and rigorous clinical studies to validate. Therefore, these findings should currently not be interpreted as an implication that obesity *per se* has a protective effect on COVID-19 outbreak. However, our findings provided timely and important evidence supporting the necessity of limited out-door exercise and Stay-At-Home policy for preventing COVID-19, and warn the public advice that obese people is more vulnerable to COVID-19 infection, which may lead to a false safety message probably given to people with normal body weight, especially during the phase of reopening.

## Data Availability

Data is available in the supplementary file

## Acknowledgement

This project is partially supported by USDA/ Hatch project (1013548-MAS00514, ZL).

## References

1. Zhu N, Zhang D, Wang W, et al. A Novel Coronavirus from Patients with Pneumonia in China, 2019. N Engl J Med. Feb 20 2020;382(8):727–733.

2. World Health Organisation. WHO Director-General’s opening remarks at the media, briefing on COVID-19 - 11 March 2020.

3. COVID-19 Dashboard by the Center for Systems Science and Engineering (CSSE) at Johns Hopkins University (JHU). ArcGIS. Johns Hopkins University. Retrieved 31 May 2020.

4. Zhong NS, Zheng BJ, Li YM, et al. Epidemiology and cause of severe acute respiratory syndrome (SARS) in Guangdong, People’s Republic of China, in February, 2003. Lancet. Oct 25 2003;362(9393):1353–1358.

5. Assiri A, McGeer A, Perl TM, et al. Hospital outbreak of Middle East respiratory syndrome coronavirus. N Engl J Med. Aug 1 2013;369(5):407–416.

6. Wang D, Hu B, Hu C, et al. Clinical Characteristics of 138 Hospitalized Patients With 2019 Novel Coronavirus-Infected Pneumonia in Wuhan, China. JAMA. Feb 7 2020.

7. Chen N, Zhou M, Dong X, et al. Epidemiological and clinical characteristics of 99 cases of 2019 novel coronavirus pneumonia in Wuhan, China: a descriptive study. Lancet. Feb 15 2020;395(10223):507–513.

8. Guan WJ, Ni ZY, Hu Y, et al. Clinical Characteristics of Coronavirus Disease 2019 in China. N Engl J Med. Apr 30 2020;382(18):1708–1720.

9. Ogden CL, Carroll MD, Curtin LR, McDowell MA, Tabak CJ, Flegal KM. Prevalence of overweight and obesity in the United States, 1999-2004. Jama. Apr 5 2006;295(13):1549–1555.

10. Flegal KM, Carroll MD, Ogden CL, Curtin LR. Prevalence and trends in obesity among US adults, 1999-2008. Jama. Jan 20 2010;303(3):235–241.

11. Silventoinen K, Sans S, Tolonen H, et al. Trends in obesity and energy supply in the WHO MONICA Project. Int J Obes Relat Metab Disord. May 2004;28(5):710–718.

12. Wang Y, Monteiro C, Popkin BM. Trends of obesity and underweight in older children and adolescents in the United States, Brazil, China, and Russia. Am J Clin Nutr. Jun 2002;75(6):971–977.

13. Calle EE, Kaaks R. Overweight, obesity and cancer: epidemiological evidence and proposed mechanisms. Nat Rev Cancer. Aug 2004;4(8):579–591.

14. Finkelstein EA, Khavjou OA, Thompson H, et al. Obesity and severe obesity forecasts through 2030. Am J Prev Med. Jun 2012;42(6):563–570.

15. Kopelman PG. Obesity as a medical problem. Nature. Apr 6 2000;404(6778):635–643.

16. Gonzalez-Muniesa P, Martinez-Gonzalez MA, Hu FB, et al. Obesity. Nat Rev Dis Primers. Jun 15 2017;3:17034.

17. Schmidt FM, Weschenfelder J, Sander C, et al. Inflammatory cytokines in general and central obesity and modulating effects of physical activity. PLoS One. 2015;10(3):e0121971.

18. Ellulu MS, Patimah I, Khaza’ai H, Rahmat A, Abed Y. Obesity and inflammation: the linking mechanism and the complications. Arch Med Sci. Jun 2017;13(4):851–863.

19. Wu Z, McGoogan JM. Characteristics of and Important Lessons From the Coronavirus Disease 2019 (COVID-19) Outbreak in China: Summary of a Report of 72314 Cases From the Chinese Center for Disease Control and Prevention. JAMA. Feb 24 2020.

20. Chiappetta S, Sharma AM, Bottino V, Stier C. COVID-19 and the role of chronic inflammation in patients with obesity. Int J Obes (Lond). May 14 2020.

21. Petrilli CM, Jones SA, Yang J, et al. Factors associated with hospital admission and critical illness among 5279 people with coronavirus disease 2019 in New York City: prospective cohort study. BMJ. May 22 2020;369:m1966.

22. Simonnet A, Chetboun M, Poissy J, et al. High prevalence of obesity in severe acute respiratory syndrome coronavirus-2 (SARS-CoV-2) requiring invasive mechanical ventilation. Obesity (Silver Spring). Apr 9 2020.

23. Kass DA, Duggal P, Cingolani O. Obesity could shift severe COVID-19 disease to younger ages. Lancet. May 16 2020;395(10236):1544–1545.

24. Lighter J, Phillips M, Hochman S, et al. Obesity in patients younger than 60 years is a risk factor for Covid-19 hospital admission. Clin Infect Dis. Apr 9 2020.

25. Petrakis D, Margina D, Tsarouhas K, et al. Obesity a risk factor for increased COVID19 prevalence, severity and lethality (Review). Mol Med Rep. Jul 2020;22(1):9–19.

26. Jose RJ, Manuel A. Does Coronavirus Disease 2019 Disprove the Obesity Paradox in Acute Respiratory Distress Syndrome? Obesity (Silver Spring). Jun 2020;28(6):1007.

27. Ni YN, Luo J, Yu H, et al. Can body mass index predict clinical outcomes for patients with acute lung injury/acute respiratory distress syndrome? A meta-analysis. Crit Care. Feb 22 2017;21(1):36.

28. Li Y, Li Q, Zhang N, Liu Z. Sunlight and vitamin D in the prevention of coronavirus disease (COVID-19) infection and mortality in the United States. Research Square. DOI: 10.21203/rs.3.rs-32499/v1. 2020.

29. Ilie PC, Stefanescu S, Smith L. The role of vitamin D in the prevention of coronavirus disease 2019 infection and mortality. Aging Clin Exp Res. May 6 2020.

30. Rhodes JM, Subramanian S, Laird E, Kenny RA. Editorial: low population mortality from COVID-19 in countries south of latitude 35 degrees North supports vitamin D as a factor determining severity. Aliment Pharmacol Ther. Jun 2020;51(12):1434–1437.

31. Ryan DH, Ravussin E, Heymsfield S. COVID 19 and the Patient with Obesity - The Editors Speak Out. Obesity (Silver Spring). May 2020;28(5):847.

32. Flint SW, Tahrani AA. COVID-19 and obesity-lack of clarity, guidance, and implications for care Lancet Diabetes Endocrinol. Jun 2020;8(6):474–475.

33. O’Brien JM, Jr., Phillips GS, Ali NA, Lucarelli M, Marsh CB, Lemeshow S. Body mass index is independently associated with hospital mortality in mechanically ventilated adults with acute lung injury. Crit Care Med. Mar 2006;34(3):738–744.

34. Bustamante AF, Repine JE. Adipose-lung cell crosstalk in the obesity-ARDS paradox. J Pulm Respir Med. 2013(3):10001444.

35. Bluher M. Obesity: global epidemiology and pathogenesis. Nat Rev Endocrinol. May 2019;15(5):288–298.

36. Wu C, Chen X, Cai Y, et al. Risk Factors Associated With Acute Respiratory Distress Syndrome and Death in Patients With Coronavirus Disease 2019 Pneumonia in Wuhan, China. JAMA Intern Med. Mar 13 2020.

37. Hotamisligil GS. Inflammation and metabolic disorders. Nature. Dec 14 2006;444(7121):860–867.

